# AI Implementation in U.S. Healthcare and Its Association With Elder Mortality and Quality of Care

**DOI:** 10.1101/2025.06.24.25330241

**Authors:** Yeon-Mi Hwang, Madelena Y. Ng, Michelle P. Sahai, Malvika Pillai, Tina Hernandez-Boussard

## Abstract

Hospitals are increasingly adopting artificial intelligence (AI) tools in clinical care. However, their overall impact on the health of older adults remains unclear. We assessed whether county-level hospital AI implementation was associated with elder mortality and care quality using national data from the American Hospital Association (AHA), Centers for Medicare & Medicaid Services (CMS), and CDC WONDER. Higher AI implementation was associated with lower elder deaths from all causes, circulatory and respiratory conditions, along with improvements in pneumonia and heart failure mortality, reduced hospital readmissions, and staff vaccination rates. However, we also found associations between AI implementation and increased rates of high-risk opioid prescribing and sepsis shock incidence. These findings highlight both the potential benefits and unintended consequences of AI in elder care. Given limitations in data specificity and the cross-sectional study design, future research should adopt longitudinal methods and promote more standardized reporting of AI use.

## Introduction

Healthcare systems increasingly use predictive analytics and artificial intelligence (AI) technologies for clinical decision-making, risk stratification, and care coordination.^1^ Despite growing implementation, evidence on whether these tools improve population health outcomes and care quality, particularly for elders, remains limited. Given the aging U.S. population and elders’ disproportionate healthcare utilization,^2^ understanding AI’s community-level impact is critical for health policy and resource allocation decisions. We examined whether county-level AI implementation in hospitals was associated with elder mortality rates and Center for Medicare & Medicaid Services (CMS) care quality indicators^3^ to understand their impact on community-level elder health and inform policy discussions on AI in healthcare delivery.

## Methods

This cross-sectional study used three data sources: the 2023 American Hospital Association (AHA) Information Technology (IT) Supplement^4^ to identify hospital AI use, the February 2025 CMS Hospital Quality of Care data for quality metrics^3^, and the 2023 Centers for Disease Control and Prevention Wide-Ranging Online Data for Epidemiologic Research (CDC WONDER) data for county-level mortality.^5^ Hospitals were classified as using no predictive models (0), non-AI predictive models (1), or AI models (2). County-level AI implementation was defined as the average score among hospitals in each county.

We assessed the association between county AI implementation level and hospital quality or elder mortality outcomes with <40% missing data using multivariate linear regression adjusting for hospital characteristics (bed size, teaching status, critical access, rural referral center, ownership, and payer mix). Mortality outcomes were specific to elders, while hospital quality metrics represented general care processes. Multiple testing was corrected using the Benjamini-Hochberg method for false discovery rate control.

## Results

Of 3220 U.S. counties, 1640 (50.9%) had at least one hospital with reported AI implementation data (Figure 1). Geographic variation in AI implementation reveals disparities in healthcare technology access across communities. Among 22 county-level outcomes with <40% missingness, 10 showed significant associations with AI implementation level (Figure 2).

**Figure 1.**
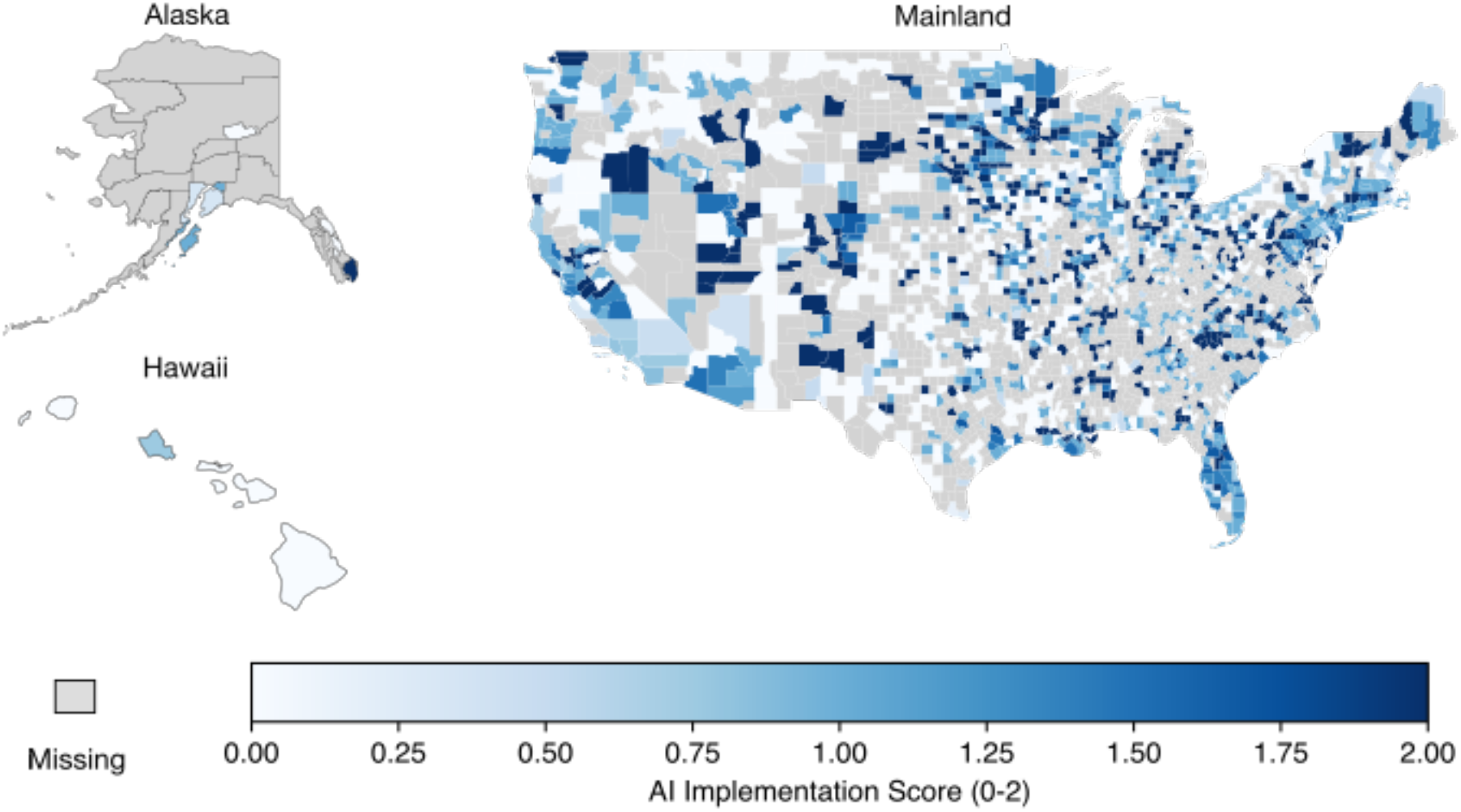
Distribution of Healthcare System AI Implementation Level Across U.S Counties. Abbreviation: AHA, American Hospital Association; AI, Artificial Intelligence; IT, Information Technology. The mean and median of county AI implementation levels were 0.89 and 1.00, respectively. Hospitals that responded to the AHA IT supplement but did not respond to the AI implementation-specific survey question were assigned a value of 0.

**Figure 2.**
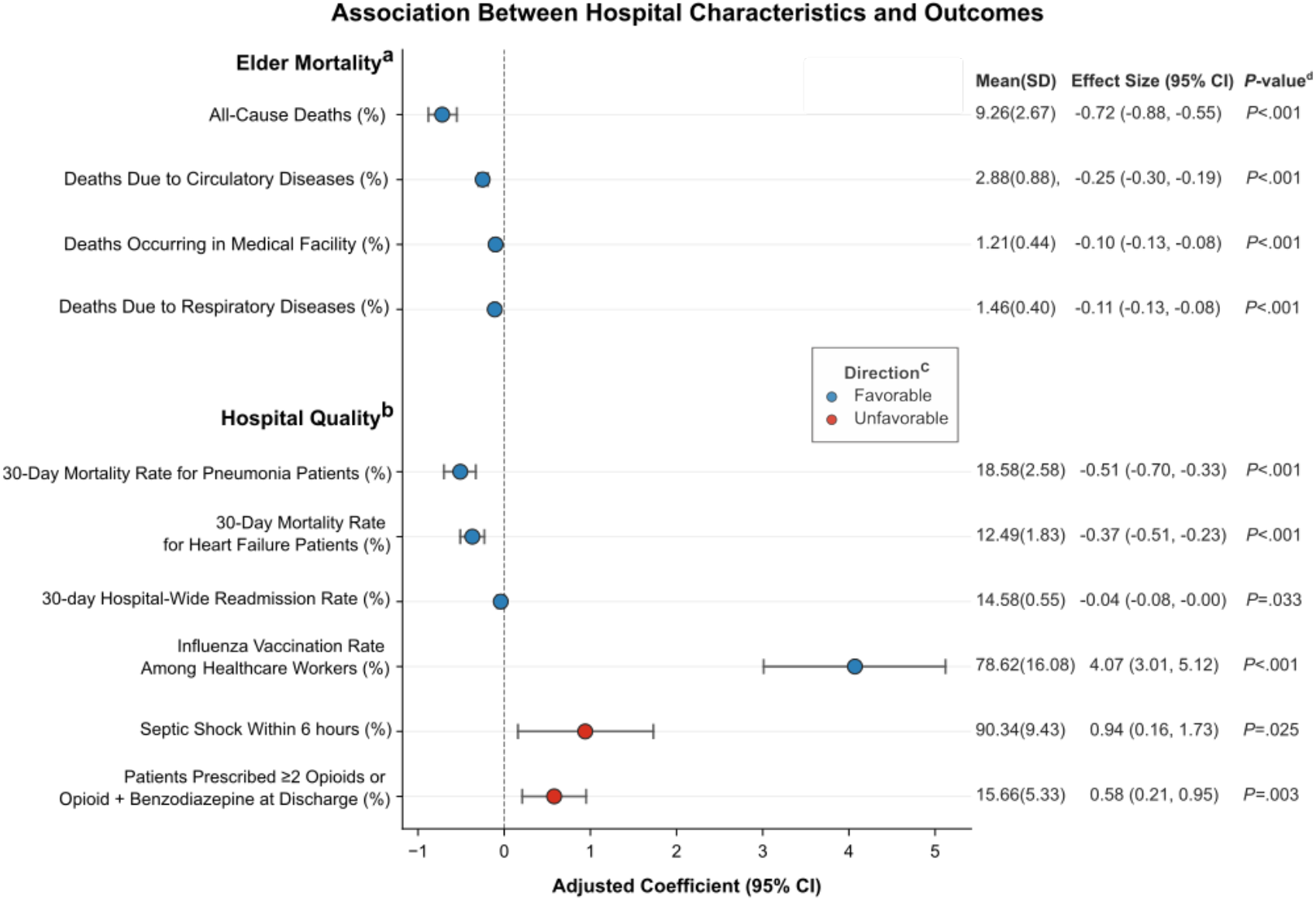
Association Between County-Level AI Implementation and Elder Health Outcomes. Abbreviation: CDC WONDER, Centers for Disease Control and Prevention Wide-Ranging Online Data for Epidemiologic Research; CMS, Center for Medicare & Medicaid Services; CI, Confidence Interval; SD, Standard Deviation. ^a^ Elder mortality rate was calculated using the 65+ death count from CDC WONDER for 2023. To account for the population aged 65+ in each county, we divided the mortality count by the 2023 population aged 65+ obtained from the U.S. Census Bureau^7^ for that specific county. ^b^ Hospital quality of care data were collected from the most recent (as of May 2, 2025) February 2025 CMS Hospital Care Compare.^3^ ^c^ Direction (favorable, unfavorable) indicates a favorable outcome (lower mortality rate or better care quality) according to standards and to guide reader interpretation. ^d^ Multiple testing was corrected using the Benjamini-Hochberg method for false discovery rate control.

For elder mortality, each one-level increase in AI implementation was significantly associated with decreases in all-cause deaths (−0.72% [−0.88, −0.55]; *P*<.001), circulatory disease deaths (−0.25% [−0.30, −0.19] *P*<.001), deaths in medical facilities (−0.10% [−0.13, −0.08]; *P*<.001), and respiratory disease deaths (−0.11% [−0.13, −0.08]; *P*<.001).

For hospital quality metrics, each one-level increase in AI implementation was significantly associated with lower 30-day pneumonia mortality (–0.51% [–0.70, –0.33]; *P*<.001), lower 30-day heart failure mortality (–0.37% [–0.51, –0.23]; *P*<.001), reduced 30-day hospital-wide readmissions (–0.04% [–0.08, 0.00]; *P*=.033), higher staff influenza vaccination (4.07% [3.01, 5.12]; *P*<.001), higher sepsis shock incidence (0.94% [0.16, 1.73]; *P*=.025), and increased discharge prescribing of ≥2 opioids or concurrent opioid and benzodiazepine (0.58% [0.21, 0.95]; *P*=.003).

## Discussion

This study provides the first national quantification of associations between hospital AI implementation and elder health outcomes. Findings highlight AI’s potential to improve patient outcomes but reveal disparities and risks. Reductions were observed for elder all-cause, circulatory, respiratory mortality, and deaths in medical facilities. Quality improvements included lower pneumonia and heart failure mortality, fewer readmissions, and higher influenza vaccination rates. However, AI implementation was also associated with increased high-risk opioid prescribing and septic shock incidence, which may reflect differences in clinical complexity, hospital protocol, or care practices. These quality metrics are directly connected to elder health, as benefits and risks we identified disproportionately affect older adults due to age-related physiological vulnerabilities.

Our findings should be interpreted with caution. The cross-sectional design prevents causal inference and our data lacks specificity regarding AI types, use case, or integration. Hospitals adopting AI often have greater financial resources, IT infrastructure, and innovation readiness.^1^ These institutional differences likely correlate with regional socioeconomic disparities that were not fully captured in our analysis.^6^ Despite these limitations, the diversity of U.S. hospital settings in our national sample provides valuable insights into AI implementation effects across different healthcare environments.

Future research should employ longitudinal designs to establish temporal relationships, explore hospital-level variation in AI implementation incorporating local context and socioeconomic factors, and examine specific AI use cases. Regulatory approaches should address the lack of standardized hospital reporting on AI implementation details, which limits nuanced analysis. Future assessments should incorporate elder-specific outcomes when evaluating these technologies. Policymakers should develop elder-focused AI guidelines that acknowledge distinct health needs of aging populations and require transparent reporting to facilitate research on equity and effectiveness for this vulnerable demographic.

## Article Information

### Author Contributions

Drs. Yeon-Mi Hwang and Madelena Y. Ng had full access to all of the data in the study and take responsibility for the integrity of the data and the accuracy of the data analysis.

*Concept and design: Hwang, Ng, Hernandez-Boussard*.

*Acquisition, analysis, or interpretation of data: Hwang, Ng*.

*Drafting of the manuscript: Hwang, Ng*.

*Critical review of the manuscript for important intellectual content: All authors*.

*Statistical analysis: Hwang*.

*Obtained funding: Ng, Hernandez-Boussard*.

*Administrative, technical, or material support: Hernandez-Boussard*.

*Supervision: Hernandez-Boussard*.

### Human Ethics and Consent to Participate declarations

Not applicable.

### Data Sharing Statement

Analysis used data from CDC WONDER, U.S. Census Bureau, and Centers for Medicare & Medicaid Services, which are publicly available through the sources cited in the references. American Hospital Association (AHA) data requires a subscription and can be obtained through AHA. Information on data access is available in the cited references.

